# Modeling COVID-19 on a network: super-spreaders, testing and containment

**DOI:** 10.1101/2020.04.30.20081828

**Authors:** Ofir Reich, Guy Shalev, Tom Kalvari

## Abstract

To model COVID-19 spread, we use an SEIR agent-based model on a graph, which takes into account several important real-life attributes of COVID-19: super-spreaders, realistic epidemiological parameters of the disease, testing and quarantine policies. We find that mass-testing is much less effective than testing the symptomatic and contact tracing, and some blend of these with social distancing is required to achieve suppression. We also find that the fat tail of the degree distribution matters a lot for epidemic growth, and many standard models do not account for this. Additionally, the average reproduction number for individuals, equivalent in many models to *R*_0_, is not an upper bound for the effective reproduction number, *R*. Even with an expectation of less than one new case per person, our model shows that exponential spread is possible. The parameter which closely predicts growth rate is the ratio between 2nd to 1st moments of the degree distribution. We provide mathematical arguments to argue that certain results of our simulations hold in more general settings.

## Summary

We use a model of COVID-19 spread, an SEIR[5] agent-based model on a graph[2], which takes into account several important real-life attributes of COVID-19:

- **Super-spreaders**[6], i.e. the fact that some people have many more connections than others, are more likely to be infected and if infectious will infect many more people than the median. We use a power-law degree distribution.
- **Realistic epidemiological parameters** of the disease:
  – Doubling time when no action is taken.
  – Durations of different stages of disease progression (incubation, symptoms, recovery).
  – Different infectiousness at each of these different stages.
  – Importantly, we do *not* rely on estimates of *R*_0_, which are very uncertain and depend on other assumptions.^1^
- **Testing and quarantine policies**.

We provide simulation results and mathematical arguments to argue that these results hold more generally. We draw conclusions both about graph structure and about testing & quarantine policies.

### Graph structure

The main conclusions about graph structure are:

- **The average reproduction number for individuals**, *r*, **equivalent in many models to** *R*_0_, **is not an upper bound for the effective reproduction number**, *R*. **Even with an expectation of less than one new case per person, our model shows that exponential spread is possible**. This is because of super-spreaders - the fat tail of the degree distribution - who are more likely to get infected, and when infectious infect many people.
- **The degree distribution (not just its mean) matters a lot** both for the epidemic growth rate, and for the eventual outcome (number of people infected). Many standard models neglect this (sometimes implicitly, using a model equivalent to constant degree, or with degree distribution with a small standard deviation). These models can produce systematic errors. Social distancing could be effective by lowering the mean degree (which matters a lot), but it could also be effective by limiting super-spreaders.
- **A potentially highly cost-effective social distancing strategy might therefore be to limit super-spreaders**, e.g. by frequent testing, by limiting their contagiousness or by reducing their number of contacts, without changing the total number of contacts in the population.
- **The parameter which closely predicts growth rate is the ratio of 2nd to 1st moments of the degree distribution**: *µ*_2_/*µ*_1_ = *E*[*D*^2^]/*E*[*D*] where *D* is the degree of a random node. If the average reproduction number for individuals is *r*, we get

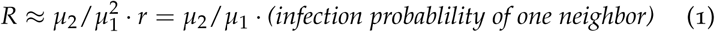
- Graph locality, the fact that neighbors of a given node tend to be themselves connected, could also be important, but we don’t model it yet. Other properties of the graph, like large highly connected and sparsely connected subgraphs, could also be important and aren’t modeled.

### Testing & quarantine

The main conclusions from testing & quarantine policy simulations are:

- **Any relaxation of lockdowns without other measures only resets the clock until the outbreak is critical**.
- **Safely exiting lockdowns depends primarily on testing capacity and tracing capacity - tests per day, swiftness of results and actions**. Without those measures driving *R* below 1, exiting lock-down would cause re-emergence of the epidemic. For limited testing capacity, a more aggressive quarantine policy can serve as a substitute.
- **Better testing, tracing & quarantine policies can create containment with far less social distancing** if they are effective enough good coverage, quick execution.
- **Without full lockdown, mass testing of the general population to search for unknown infected individuals is mostly futile for containment**, since it would require near universal testing to be effective, which is far beyond current capacity. Limited testing resources are much better spent testing the symptomatic and their contacts.

### Introduction & Model Specification

We use a Susceptible-Exposed-Infected-Recovered (SEIR) model[5] for disease progression. We use an agent-based model, where each agent (person) is represented as a node in a graph, and infection happens between contacts, represented by graph edges.^2^ We also model testing and a Quarantine group of those who test positive, and contact tracing of their neighbors in the graph. Each node belongs to exactly one of the following groups:

**Susceptible (S)**: Nodes which weren’t infected yet. These are the only nodes which can be infected. When infected, they become Exposed.

**Figure 1:**
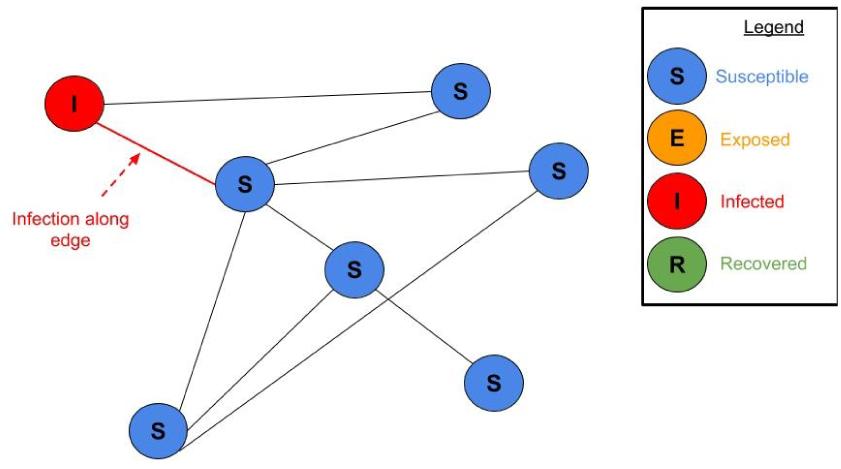
Infected nodes can infect neighboring Susceptible nodes - those who share an edge with them. Full ani-mation at bit.ly/seir-graph-animation.
**Exposed (E)**: Nodes which were infected and are now in their pre-symptomatic incubation period. They are infectious starting 2 days before the end of their (Gamma distributed) incubation period, at which point they develop symptoms (and become Infected).
**Infected (I)**: Nodes which are now symptomatic. More infectious than during the final Exposed period. When their (exponentially distributed) Infected period ends, they become Recovered.
**Recovered (R)**: Nodes who have recovered (or died). They are no longer infectious and cannot be infected.

In addition to those groups, a node can also belong to additional groups:

**Tested Positive (TP)**: Nodes who tested positive for COVID-19. At each simulation step, a certain share of Infected nodes are tested and a certain share of all nodes are tested. Nodes which are Exposed or Infected test positive. They become Quarantined, and potentially their neighbors are as well. We do not model false positives.
**Quarantined (Q)**: Nodes which are in quarantine. They can neither infect nor get infected. Quarantine ends after a set period. Their disease progression continues irrespective of Quarantine.

We take as many parameters for the model as we can find from real attributes of covid-19, and calibrate the total infection probability to produce the doubling time of 3.1 days observed in various regions. For more details about the model, see appendix below, and open source code.

Simulation starts with a certain number of Infected and Exposed nodes, and simulates the progression, tracking the total number of nodes in each group for later analysis. The results of a single simulation are plotted in the figure here. In this example there are no quarantines at all, and the epidemic spreads exponentially. In the early days, the number of Exposed outnumbers the Infected. The epidemic spreads until herd immunity is reached, at which point it decays exponentially until it is eradicated. The eventual number of people who were infected is the final number of Recovered, about 60%.

### Graph Structure Dramatically Affects Epidemic Growth

Our model can run on any graph. To model super-spreaders, we use here a power law degree distribution for nodes, which is a linear transformation of *X = U^−γ^*, where *U ~ U*[0, 1] is distributed uniformly between 0 and 1. The degree is *D* = *aX + b* so that the minimum value of *D* is 2 and the mean value of *D* is a parameter we vary. Nodes are connected to each other to satisfy the degree constraint, but otherwise randomly, so there is no locality.

Gamma is important - the higher gamma is, the fatter the tail of the degree distribution, so there are more super-spreaders with stronger spreading. The power law distribution has a fatter tail than a normal or an exponential distribution. The degree distribution in log scale is in the figure on the right.

**Figure 2:**
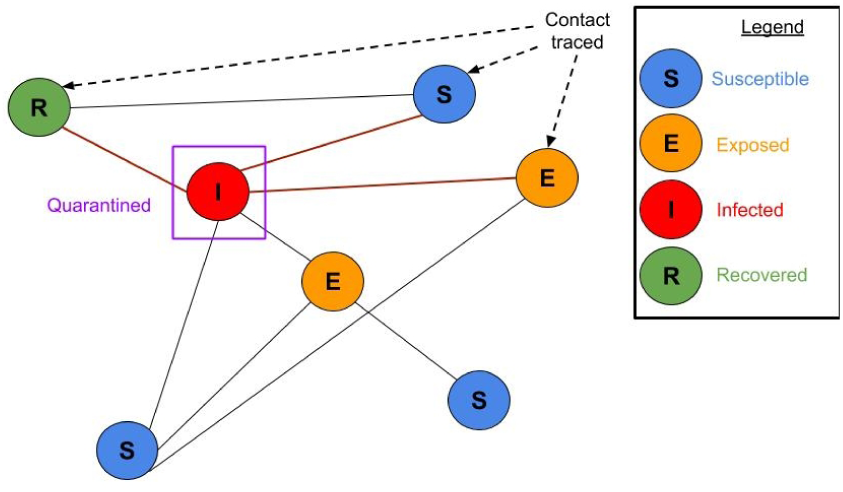
Contact tracing. Some share of the neighbors of a node which tests positive are traced and tested themselves.

**Figure 3:**
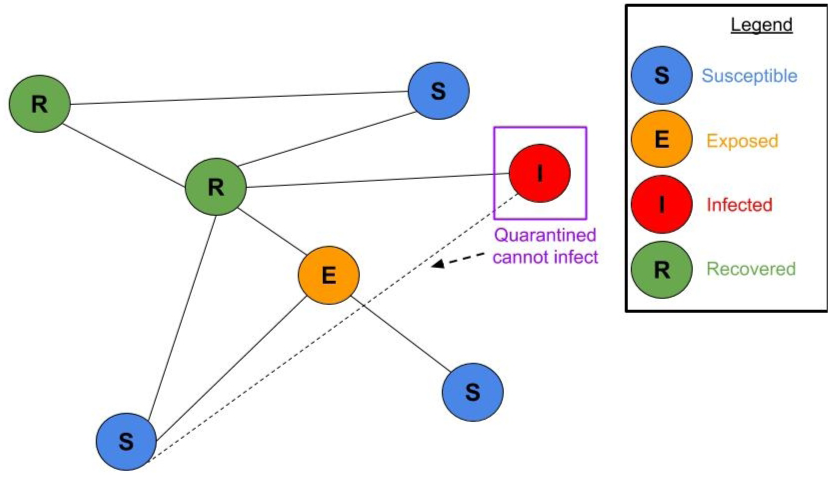
Quarantined node cannot infect others. Susceptible, Exposed, Infected and Recovered nodes shown.

**Figure 4:**
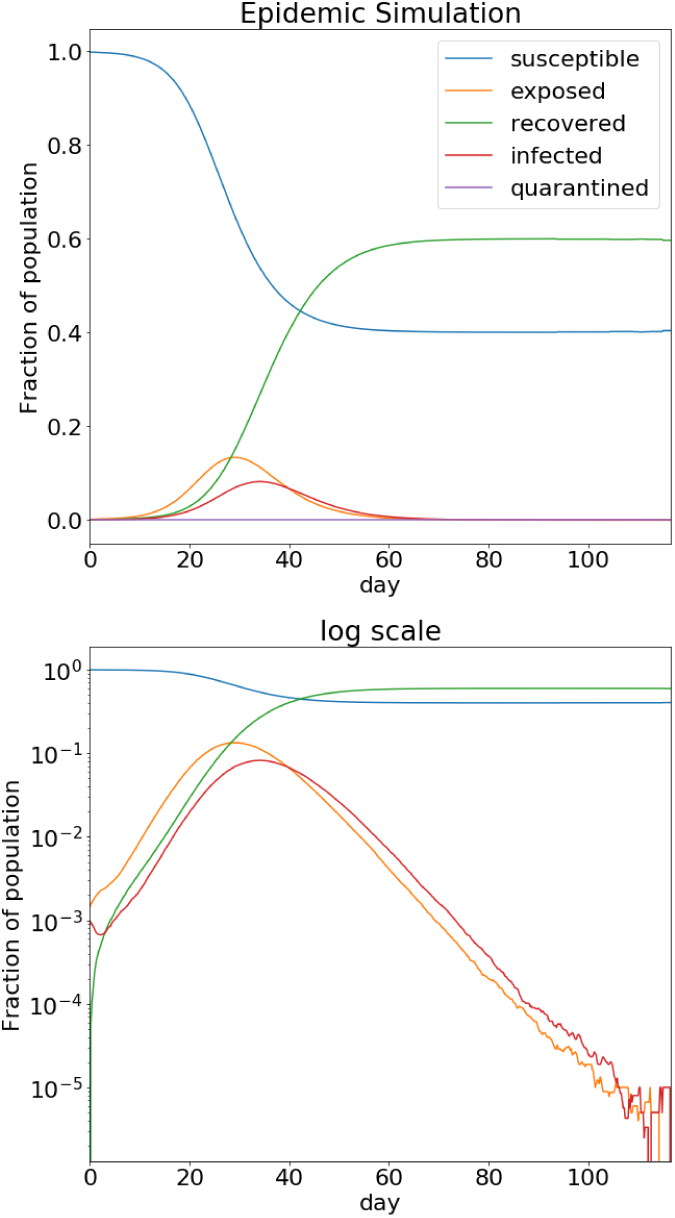
Epidemic Simulation. The X axis is time and the Y axis is the fraction of the population in each group in linear scale (top) and log scale (bottom).

### The average reproduction number, r

We define the average reproduction number, *r*, as the expected number of individuals infected by a single random person if that person is infectious and all others are Susceptible. More precisely the definition is:

- Select a node in the graph at random, uniformly, so that each node has an equal probability of being selected.
- Assume this node was infected (so it becomes Exposed, Infected, then Recovered), but all other nodes are Susceptible. Let *r_node_* be the expected number of its neighbors that it would infect until it recovers.
- *r* is the unweighted average of *r_node_* over all nodes. 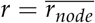.

For example, assume half of the nodes have degree 10 and half have degree 30, the disease lasts exactly 1 day and the daily probability of infecting a neighbor is 0.1, then *r_node_* = 10 · 1 · 0.1 = 1 for half of the nodes, and *r_node_* = 30 · 1 · 0.1 = 3 for the other half. So 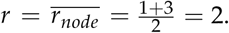

The advantage of this definition is that it is unambiguous and can be calculated precisely from the properties of the graph and the epidemiological parameters. This definition is very closely related to *R*_0_. The only difference is the uniform selection of the initial node, which is crucial as we shall see. *r* is nearly proportional to the mean degree times the probability of infection. For a homogeneous population, where the degree of all nodes is constant, as in many models, we have *R*_0_ = *r*. Therefore *r* is the reproduction number effectively used in many models and definitions. We will show it is systematically wrong as an approximation for *R*_0_.

### Degree distribution, super-spreaders and growth rate

For a specific gamma value, *r* is a good determinant of growth rate. But higher gamma values give a higher growth rate for the same *r* and the same mean degree. This is through a fatter tail of the degree distribution - super-spreaders. This causes R (the expected number of people that an infected node in the simulation infects) to be greater than *r* (the expected number of nodes that a uniformly random node would infect). As a specific case, there are many cases where *r* is less than one, but we get exponential growth in the number of infected individuals, sometimes very rapid growth.

**Figure 5:**
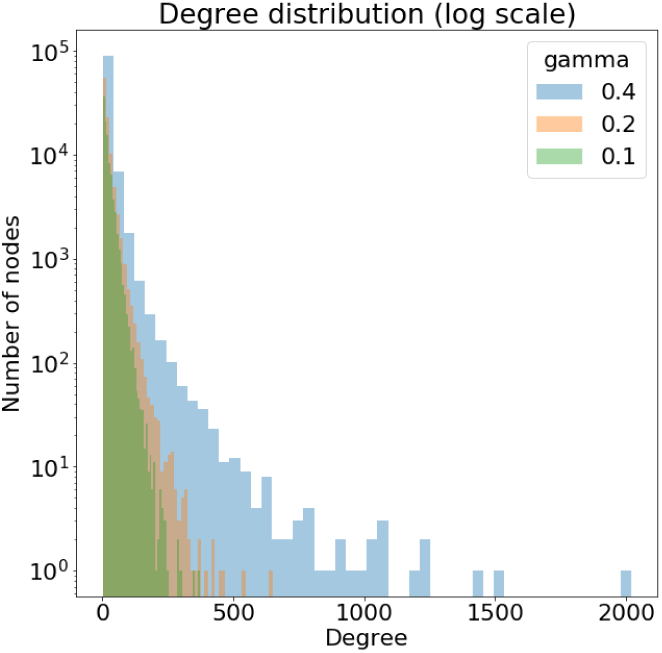
Different degree distributions corresponding to different gamma values. Higher gamma values give a fatter tail to the distribution. Here *mean degree* =20, *N* =10^5^ nodes.

In the graph on the right each point represents a simulation with certain parameters (in fact, 10 simulations which are averaged). We varied the mean degree, the infection probability of a single interaction, and the degree distribution parameter gamma (0.4, 0.2, 0.1 and 0). We plot the growth rate observed in the simulation vs. *r* which can be calculated from the mean degree and the epidemiological parameters (infection probability, recovery time, etc.). The daily growth rate of an epidemic is by what factor the number of infected people grows each day. A growth rate of 2 means the number doubles each day, a growth rate of 0.5 means it halves each day. The growth rate can be easily derived from the doubling time. For example, in one simulation in this graph, for *γ* = 0.4 and *r* = 0.85 we have a daily growth rate of 1.2.

We plot a few examples of epidemic simulations, all with *r* = 0.5, the same mean degree and infection probability, but different values of gamma. The different gammas radically change the course of the epidemic. Gamma of 0.4 gives standard exponential growth until herd immunity. Gamma of 0.2 and 0.1 give a constant simmer which eventually dies out, and gamma of 0 gives quick containment.

### Standard models neglect the degree distribution

The fact that the degree distribution dramatically affects the epidemic growth rate, and effective reproduction number, is very important in practice, since many models don’t account for this fact. We review a few families of models which do not model degree distribution:

- Compartmental models which are not agent-based, like SIR models[5] where the model state is only the fraction of the population in each group, implicitly assume that each infectious node causes the same number of infections, and that each susceptible node is equally likely to be infected. This amounts to assuming constant degree, and no locality.
- Branching process models make the same modeling choice equivalent to constant degree with no locality. Even well crafted models with other strengths.[4]
- Two-dimensional (or in fact n-dimensional) spatial models, where an interaction happens if two nodes are in some proximity to each other. In this case, the degree is not constant, but it has a relatively small standard deviation - it certainly isn’t fat-tailed like a power-law distribution. Two examples of this are Paul Romer's covid-19 model and the instructional model by [3-blue-1-brown](https://www.youtube.com/watch?v=gxAaO2rsdIs).
- Even agent based models which account for many traits of the population, such as [FRED](https://fred.publichealth.pitt.edu/)[3], do not necessarily account for the degree distribution. They do, however, model locality very well.

**Figure 6:**
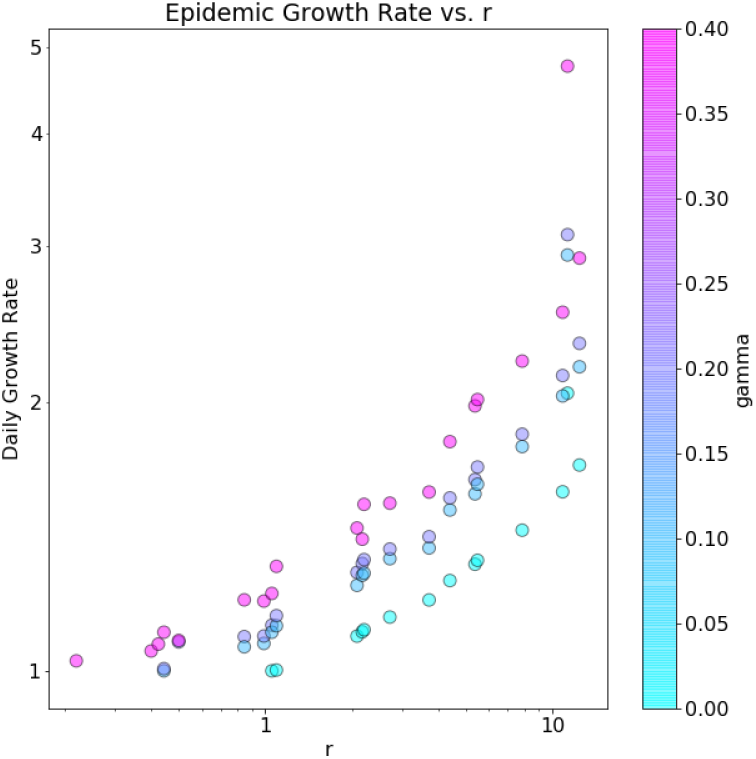
Epidemic growth rate vs. *r*, for different values of gamma (color). Broadly, higher *r* yields higher growth rate, but there is considerable hetero-geneity with higher gamma also giving higher growth rate.

**Figure 7:**
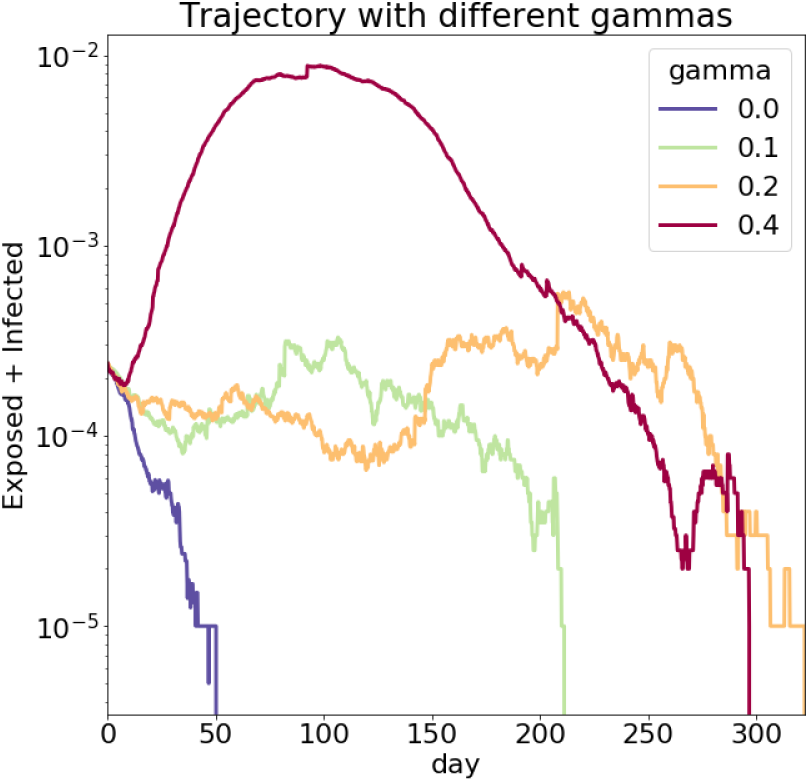
Epidemic simulations with different gamma values, plotting over time the (log) share of Exposed and Infected individuals in the population.

### A mathematical argument for 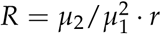

There is a simple argument for why in our case the effective reproduction number *R* before the epidemic peaks is typically greater than the average infection rate *r*. Recall our definition of *r* as the expected number of individuals infected by a single random person if that person is infectious and all others are Susceptible. This is a good approximation for the first step of infection. But after the first step of infection, individuals with larger degrees are more likely to have been infected, since they are more likely to have interacted with an infectious individual. This means after the first step, the degree distribution of the infectious is not the same as the population distribution - it skews higher. Those higher degree individuals will then infect (in expectation) more than *r* individuals, and so after the first step, *R* will be greater than *r*. An extreme (and unrealistic) case of this is a network with one central node and N-1 other nodes which are only connected to the central node. Let’s take infection with probability 1 for simplicity. In this case *r* ≈ 2, but the expected number of individuals infected in 2 steps is N-1, all other nodes, and much more than *r*^2^, which is the expected in two steps if *R* = *r*.

More precisely, if the degree, *D*, in the population is distributed by *P*(*d*)= *P*_0_(*d*), then irrespective of the degree distribution of the infectious nodes in the first generation, in the second generation it will be *P*_1_(*d*)= *C* · *P*_0_(*d*) · *d*, by Bayes’ theorem, because a node with degree *d* is *d* times more likely to be connected to an infectious node than a node with degree 1 is.^3^ Since these probabilities must sum up to 1, the constant *C* comes out *C* = 1/ ∑*_d_P*_0_(*d*) · *d* = 1/*E*[*D*]. This assumes independence between a node’s degree and its neighbors’ degrees (or infectiousness status). So we have

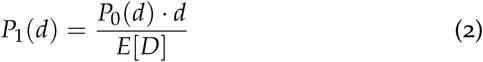

Suppose we start with one random infected node. That node has expected degree *E*[*D*] and will infect *r* nodes (in expectation, by definition of r). Those infected nodes have a higher mean degree.

What is their expected degree, *D*_1_?

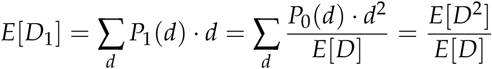

Now, these new infected nodes which have a higher mean degree will infect more than *r* nodes each. How many? If a node with degree *E*[*D*] infects *r* nodes (So *r*/*E*[*D*] per neighbor), then, since infection of each neighbor is independent of the infector’s degree, they will each infect proportionally to their degree^4^:

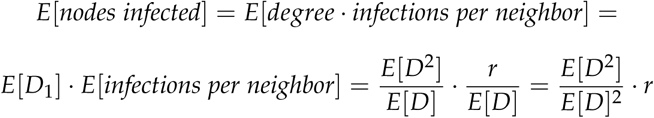

This degree distribution of infected nodes will remain roughly the same for a few steps, until Susceptible high degree nodes are exhausted. During that time, each step each infected node will cause the infection of (in expectation) 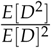 · *r* nodes, so we have

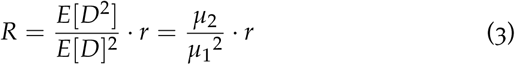

Note that if the degree is a constant *d*, this gives *R* = *d*^2^/*d*^2^ · *r* = *r* = *R*_0_ as expected.

Since *r* is proportional to the mean degree, *r* = *µ*_1_ · *P*(*infect single neighbor*), we get 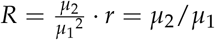 · *P*(*infect single neighbor*).

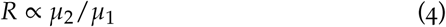

So the interesting factor is the ratio of the second to first moment *µ*_2_/*µ*_1_ of the degree distribution.^5^

We now test that this holds in our model. The daily growth rate is a linear function of *R*, which relates to the time it takes the infected node to incubate, infect and then recover (so called the “serial interval”). We therefore try predicting the growth rate rather than the (not easily observed) *R*.

On the right, we see that our theoretic *R* with the moment ratio is a far better predictor of the growth rate than *r* that we tried before. We see a robust power-law dependence, manifested as a straight line in the log-log axes, with slope 0.4. This holds for any non-local graph structure and a wide range of infection rates, whereas using *R*_0_ = *r*, as many models do, is a far worse predictor.

**Figure 8:**
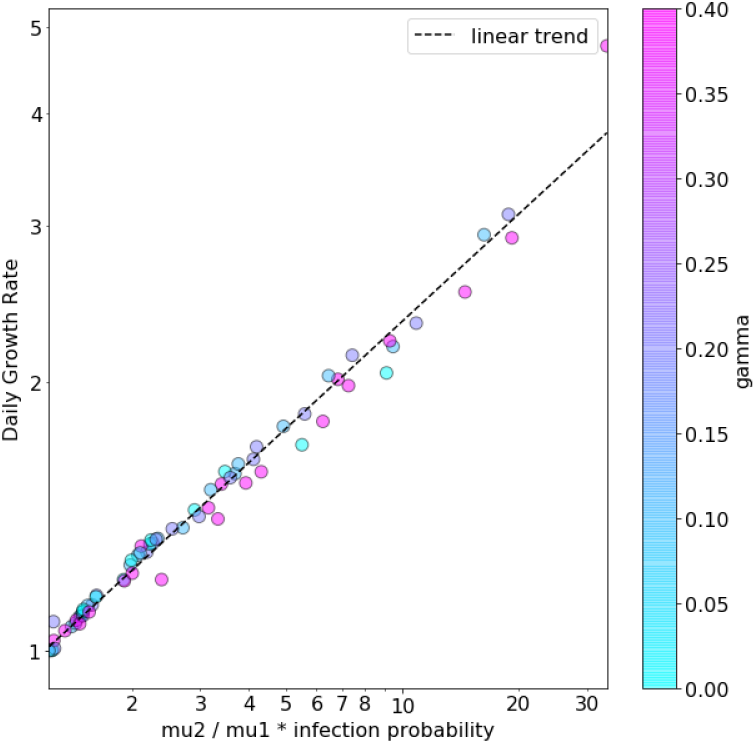
Epidemic growth rate vs. our theoretic predictor, *µ*_2_/*µ*_1_ · *infection probability*. Each point is a simulation, with its observed growth rate (y) and calculated predictor (x), for different values of gamma (color). Unlike with *r*, now all gamma values, correspond to a single straight line.

### Locality and clustering coefficient

Locality, i.e. the tendency of nodes to cluster so that neighboring nodes share many of their neighbors, is a realistic feature of human interactions, and can be very important for epidemic spread as well. Locality means an infected node has fewer nodes to infect than their degree suggests, since some of the node’s neighbors have likely already been infected by the same source which infected this node itself. The extreme version of this is if every household is isolated, where the epidemic can only spread within infected households, and then die out.

We don’t model this, but getting a more realistic graph of human interactions can yield more conclusions^6^, and this is an important avenue for future work. Note that even simulations which are very realistic and use real statistical data about schools, work places etc. still don’t always model a fat tailed degree distribution, so these are somewhat complementary approaches.

### Selecting graph parameters

We don’t have direct evidence of the true distribution of degrees (or infectiousness) in the population. There is some arbitrariness in selecting the mean degree and the infection probability. Absent direct evidence, in what follows we select gamma=0.2, which makes the degree of the top 0.1 percentile (1/1,000 nodes) equal about 15 times the median degree. We think this is a somewhat reasonable approximation of the true distribution. We select the mean degree arbitrarily to be 20, and calibrate the infection probability accordingly to produce a doubling time of 3.1 days. This is the real-life doubling time absent any action, as observed for example in US death rates.^7^

### Conclusion

Graph structure, i.e. the network of who can infect whom, has a decisive effect on the growth of epidemics. Specifically, super-spreaders, or the fat tail of the degree distribution, matter a lot, in addition to the mean degree. The expected number of infections of a random individual, *r*, in many models equivalent to *R*_0_, is not a good enough indicator of epidemic spread. It is also obviously not a property of a virus itself, since it depends on many other conditions, such as social, living & hygiene conditions. We showed that the graph structure has a very large influence on the effective reproduction number, *R*, even when *r* stays constant, because of dynamics of the epidemic spread. It is therefore crucial to model the graph structure to reach the right conclusions about epidemic spread.

## Testing & Quarantine Policies

### Results

- Exiting lockdowns without allowing critical levels of spread is possible if testing and tracing capacity are available. Without those measures driving R below 1, exiting lockdown would cause reemergence of the epidemic.
- Testing & quarantine policies can create containment even without social distancing if they are effective enough (good coverage, quick execution), or if they are less effective but combined with some social distancing.
  – Testing the general population (mass testing) is mostly futile for containment. In order to actually create containment, it needs to be done at such a high rate that requires infeasible resources. Spending the same amount of tests on symptomatic patients and their contacts is much more efficient.
  – Testing just the symptomatic can create containment, but only if it’s done with such speed and certainty that are not really feasible.
  – Tracing the contacts of those who tested positive and testing them, is another important tool for containment. And its effects might even be understated in our results, due to certain abstractions in our model, most of all lack of locality of the graph. Combined with testing the symptomatic, it can achieve containment more easily.
  – Absent test kits, such as in developing countries, diagnosing by symptoms and then aggressively tracing contacts and quarantining them without testing can be a substitute for testing.

### Outcomes of interest

To compare different policies against each other, we need to explain what the policies are and what outcomes we care about. Let’s start with the outcomes. The three main outcomes we’ll track here are the following.

- **Share of the population eventually infected**. This is the proportion of individuals who are infected at some point before the epidemic is eradicated. Some fraction of this is the deaths, so this is obviously important. We term a share of fewer than 5% “containment”.
- **Peak daily test rate**. This is the maximum number of tests performed in a single day during the course of the epidemic. It’s important because this is how we know if we can even support a certain policy, in terms of the amount of test kits it requires.
- **Quarantine days per person**. The total quarantine time experienced by all individuals throughout the epidemic, divided by the total number of individuals in the population. This gives the number of days the average person will spend in quarantine.

Keep in mind that our simulations start with 0.1% of the population Infected, and 0.15% of the population Exposed. So outcomes should sometimes be scaled to the share of the population infected when the policy is implemented, for example peak test rate.

### Policies

Our policies are divided into three main categories, according to who is tested:

- **Mass Testing - testing the general population**. A random subset of the general population (that has not tested positive before) is tested. Exposed or Infected individuals who are tested, test positive. We parameterize this by the fraction of the population tested each day.
- **Test only the symptomatic patients**, and nobody else. However, not all of them are tested, and not immediately. We model this as a random subset of Infected individuals being tested every day (and found positive). We parameterize by what fraction of the Infected are tested per day, or equivalently by the expected number of days between developing symptoms and being tested. This could also be testing not with kits, but by symptoms, for example in developing countries.
- **Test & trace**. This is a complement to testing the symptomatic or the general population, in which for those who test positive, their contacts are also tested with some probability, to find other Exposed or Infected individuals. All the positives are quarantined. We parameterize this by the share of contacts traced and tested. We also briefly consider the case where contacts aren’t tested with test kits, but instead all quarantined.

All of these policies can be combined with social distancing. We now go over these policies and report the outcomes they produce.

### Mass testing

How many tests of the general population would we have to perform every day in order to achieve containment? We’ll look at different fractions of the population tested every day (30%, 10%, 1% etc.), and graph main outcomes that result from each intensity of this policy. To create containment, we need to test 30% of the population every day. If we only test 10% of the population every day, we get 34% of the population infected - no containment (blue bars). Of course the test rate is very high in both of these options.

There’s a good argument for why it’s hard to achieve containment this way. Testing a random X% of the population per day and quarantining the positives only reduces the growth rate of the epidemic by X% a day, since it “eliminates” X% of the carriers. If the epidemic grows faster than X% a day (as in our case), this doesn’t get the growth rate below 1 and does not achieve containment. This is not to say mass testing is not effective at all, just that it requires so many tests to be effective, that these testing efforts are much better spent elsewhere. Other methods to detect carriers must be considered.

### Testing the symptomatic

Now we review policies of testing the symptomatic. Our parameter is the expected number of days between a patient developing symptoms (becoming Infected in our model) and being tested. On the right is a chart of the outcomes.

Notice if it takes 0.2 days (almost immediately) to detect a symptomatic patient and those who tested positive are immediately quarantined, we get about 2% of the population eventually infected substantial, but still containment. In this case, we need to test 0.1% of the population at the peak day. If it takes 1 day to detect a symptomatic patient, it will make 20% of the population get infected - no containment. We would also need more tests at the peak in this case, and require more quarantine time.

In other words, quarantining all the Infected immediately as they become Infected (before they infect anybody else) can stop the spread, but if it takes even one day on average to find and quarantine the symptomatic, we get exponential growth. This means even if all symptomatic patients self-isolate and stop infecting after one day of symptoms, we would still not achieve suppression. Though it does slow down the doubling time significantly, from 3.1 to 7.7 days (not shown on the chart).

The reason it’s hard for testing the symptomatic alone to create containment is that there are pre-symptomatic infections in the incubation period (the Exposed stage in our model). These infections cannot be stopped by testing the symptomatic, no matter how immediate the test is. These could be very significant for COVID-19. If these pre-symptomatic infections alone create an effective reproductive number *R* > 1, testing the symptomatic will not create containment. In our model, they create *R* of almost 1, so that even only a minor additional delay in testing is enough to allow additional infections in the early Infected period, and push *R* above 1. Something more is required.

**Figure 9:**
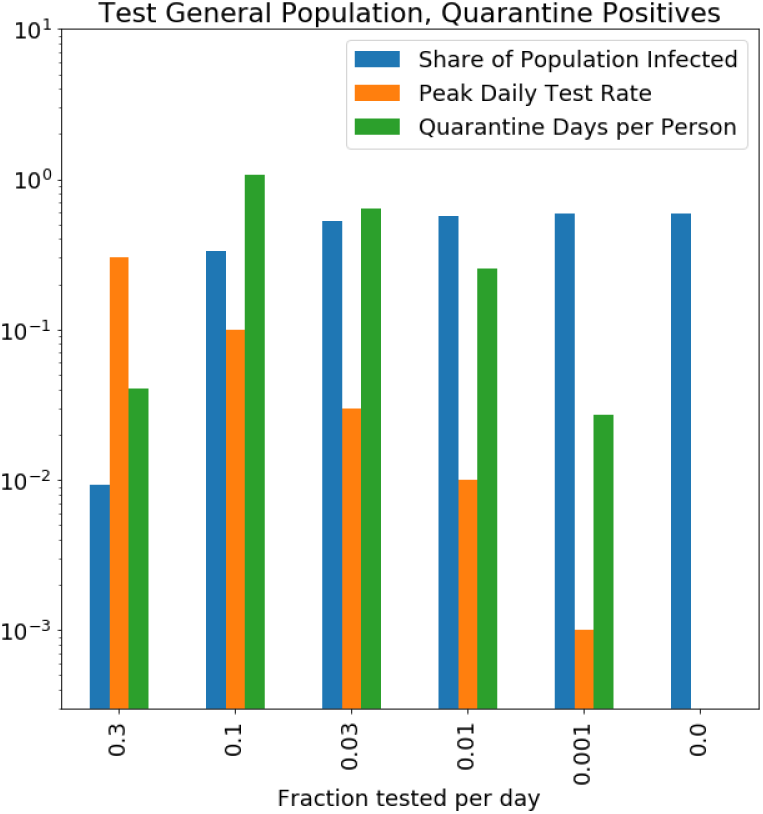
Policy outcomes of mass-testing. The X axis denotes the fraction of the population tested every day, in descending order - to the left is more testing. The Y axis tracks our 3 main outcomes (on a logarithmic scale).

**Figure 10:**
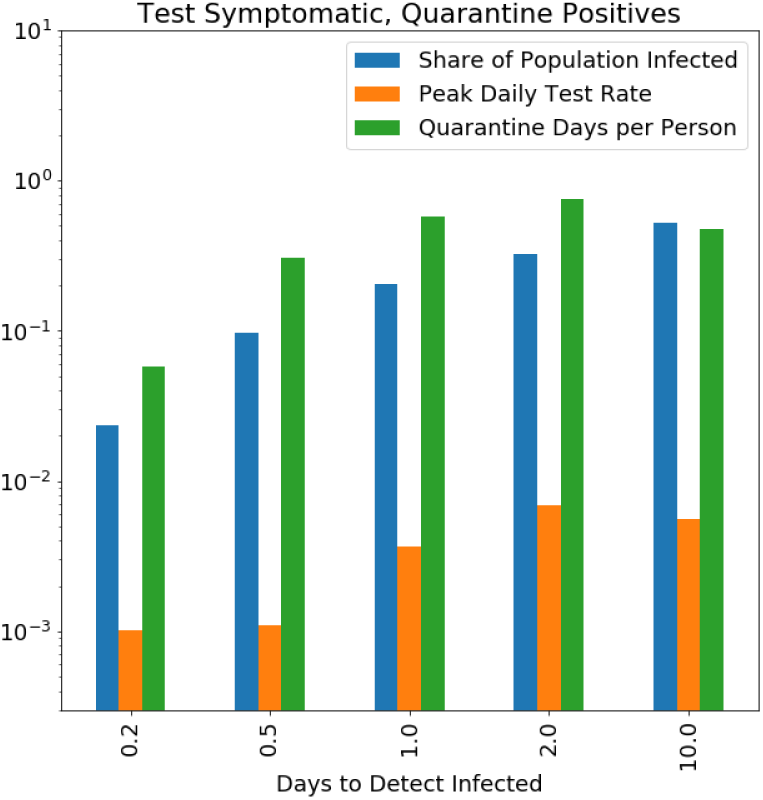
Policy outcomes of testing the symptomatic. The X axis denotes the average number of days to detect an Infected individual - left is quicker detection - and the Y axis our 3 main outcomes, again on a logarithmic scale.

### Test & trace

Quarantining the Infected is useful for containment, as we’ve seen. A way to leverage this to find more Infected and Exposed individuals is to trace the contacts of the Infected individual, and to test them as well to see if they have already been infected. This is called “test & trace”. We implemented only one cycle of this:

1. Testing of Infected / general population (with some probability, as before).
2. For those who tested positive, some fraction of their neighbors is traced and tested. Of those tested, both Infected and Exposed test positive.
3. Quarantining all those who tested positive from steps 1 and 2.

We did not model an additional steps of testing the neighbors of positives from step 2, their neighbors, and so on.

Suppose it takes 2 days on average to detect an Infected individual. The chart on the right shows how the fraction of contacts successfully traced changes our outcomes. This has a large effect. Tracing all contacts of an Infected individual who tests positive achieves containment, with 3% of the population eventually infected. Tracing only 10% of the contacts (rightmost bars) does not achieve containment, with 24% of the population eventually infected.

We think this actually understates the real life effect of test & trace. Our graph is not local at all, so there are no clusters of infected nodes. This means that neighbors of an Infected node X, which tested positive, are only more likely to be Exposed or Infected themselves to the extent that they were infected by X. They don’t have any larger chance of having been infected by the same node which infected X. In reality, there is much clustering, and so tracing can help detect these pockets of infections, and be even more effective.

**Figure 11:**
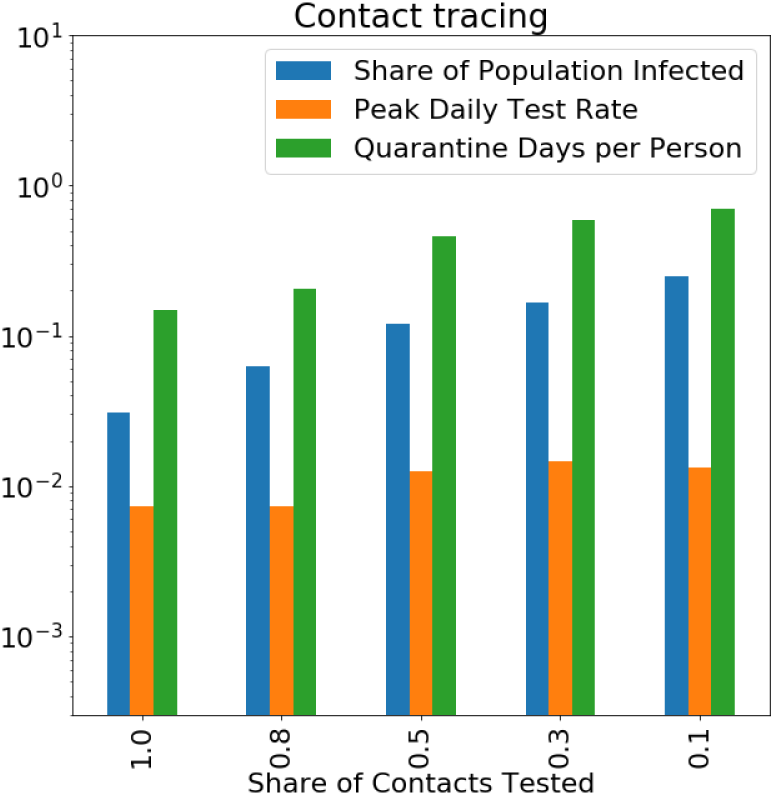
Policy outcomes of contact tracing. The X axis denotes the average share of contacts of a positive case which are traced & tested. The Y axis is the outcomes. Assumes testing the symptomatic yields detection in (on average) 2 days.

Test & trace can also be combined with mass-testing: tracing the contacts of those who test positive. Even with 100% of contacts traced and tested, still mass-testing of just over 10% of the population daily is required for containment. The peak test rate in this case is predictably very large, almost identical to mass-testing without contact tracing. Again, we see that there is not much point in mass testing.

### Limited testing capacity and more aggressive quarantine

If not enough test kits are available to test all contacts, as in developing countries, tracing might still be possible. In this case, instead of testing contacts, a more aggressive policy of quarantining all of the traced contacts without testing them can achieve results which are at least as good at containment, at the price of much more quarantine time. In a sense, testing is a substitute for more quarantine. If there is slack test capacity, where the constraint is just the cost of a single test and not the amount of tests performed per day, testing can be a way to minimize quarantine time.

### With social distancing

All of these analyses were done assuming there is no social distancing, so absent any testing & quarantines the doubling time is 3.1 days. But what if social distancing & hygiene push the infection rate down a bit? We performed all of the same simulations with a lower infection probability (about 2/3 of the original), which gives a basic doubling time of about 5 days. This does not correspond to lockdown, but to some social distancing measures still in place, say limiting large gatherings. The policies’ relative effectiveness stays the same, except that now slightly less action is required.

### Conclusion

In conclusion, there are a few tools at our disposal to decrease the effective reproduction number *R* below 1 and achieve containment:

- Testing symptomatic individuals - helps a lot, but not enough on its own. The faster and more hermetic it is, the more effective it is.
- Contact tracing - helps a lot as well.
- Social distancing to slow the base rate of spread.
- Mass testing - requires a lot of tests to have an effect, so less effective.

| Policy | Containment with<br>business as usual | Containment with some social<br>distancing (doubling time $\sim 5$ days) |
| --- | --- | --- |
| **Mass testing** |  |  |
| without contact tracing | Test 30% daily. | Test $>10\%$ daily. |
| with contact tracing | Test $>10\%$ daily,<br>trace 100% of contacts. | Test 10% daily,<br>trace 50% of contacts. |
| **Testing the symptomatic** |  |  |
| without contact tracing | Detect Infected in 0.2 day | Detect Infected in 1 day. |
| with contact tracing | Detect Infected in 1 day,<br>trace 50% of contacts. | Detect Infected in 2 days,<br>trace 50% of contacts. |

Enough of these need to be combined to drop *R* below 1. Different combinations are possible according to what is feasible and least costly. Our simulations suggest some social distancing (short of lock-down), testing of symptomatics and contact tracing are the way to go. But this is sensitive to economic calculations of costs, and the available technology. Nevertheless, these measures, or others, must be applied in order to exit lockdown without the epidemic spreading very quickly again.

## Data Availability

All data are included and referenced in the paper. The code will become publicly available very shortly on GitHub.

https://github.com/ofir-reich/seir-graph

## Appendices

### Model specification

#### Model mechanics

In more detail, here are the model mechanics. For a full specification, refer to the open source code. Each simulation step is composed of several steps which change the different groups above. See illustrative animation at bit.ly/seir-graph-animation.

> `for each simulation step:`
>
> `S, E = infection_step()`
>
> `E, I = incubation_step()`
>
> `TP = testing_step()`
>
> `Q = quarantine_step()`
>
> `I, R = recovery_step()`

**infection_step (S - > E):**

Each non-Quarantined Infected or Exposed node (in the final 2 days before becoming Infected) is infectious.
Each infectious nodes infects each of their non-Quarantined Susceptible neighbors with a certain probability.
Nodes who were infected become Exposed and start their incubation time, which has a Gamma distribution.

**incubation_step (E - > I):**

Exposed nodes whose incubation time has ended become Infected.
Exposed nodes who have less than 2 days left to become Infected become infectious, but to a lesser degree.

**testing_step (update TP):**

A test is positive if a node is either Exposed or Infectious.
Tests of symptomatic: some proportion of Infected nodes are tested (and test positive).
Mass-testing: some proportion of the entire population is tested.
Contact tracing: some proportion of neighbors of those who tested positive, are also tested.

**quarantine_step (update Q):**

Nodes who tested positive enter Quarantine for 14 days. Optionally, all neighbors of nodes who tested positive also enter Quarantine for 14 days.
Quarantined nodes who finished their quarantine time exit quarantine (and retain their SEIR state).
Quarantined nodes who tested positive in the past and are now Recovered exit Quarantine, even if their 14 days aren’t up.
Quarantined nodes who tested positive in the past and haven’t recovered stay in Quarantine, even if their 14 days are up.
Quarantined individuals can neither infect nor be infected.

**recovery_step (I - > R):**

Some proportion of Infected nodes become Recovered, and are no longer infectious. This has no memory, so it creates an exponentially distributed recovery time.

**Initial conditions (initialize all groups)**

Some fraction of nodes are randomly selected to start as Infected.
A slightly larger fraction are randomly selected to start as Exposed. Their incubation times are drawn from the same Gamma distribution of incubation time.
All other nodes start as Susceptible.
No nodes start as Recovered, Quarantined or Tested Positive.

The simulation makes several steps per day, with step-wise probabilities calculated as the daily probabilities divided by the number of steps per day. Full parameter table is below.

#### Simulation results

We typically run a simulation of the progression of the epidemic and collect aggregate stats at each simulation step: total number of Susceptible, Exposed, Infected, Recovered, Quarantined, tests performed. This creates several time series. We then typically run 10 simulations with the same model parameters and average these time series to reduce noise (see figure on the right). We calculate our final outcome stats on this average time series. There is some noise or initial condition dependence in the individual simulations, which is typically smoothed out in the average. Our simulation outcomes can include the total number of nodes infected, the peak test rate, the total quarantine time, the doubling time during the exponential spread and more.

#### Strengths and limitations

**Figure 12:**
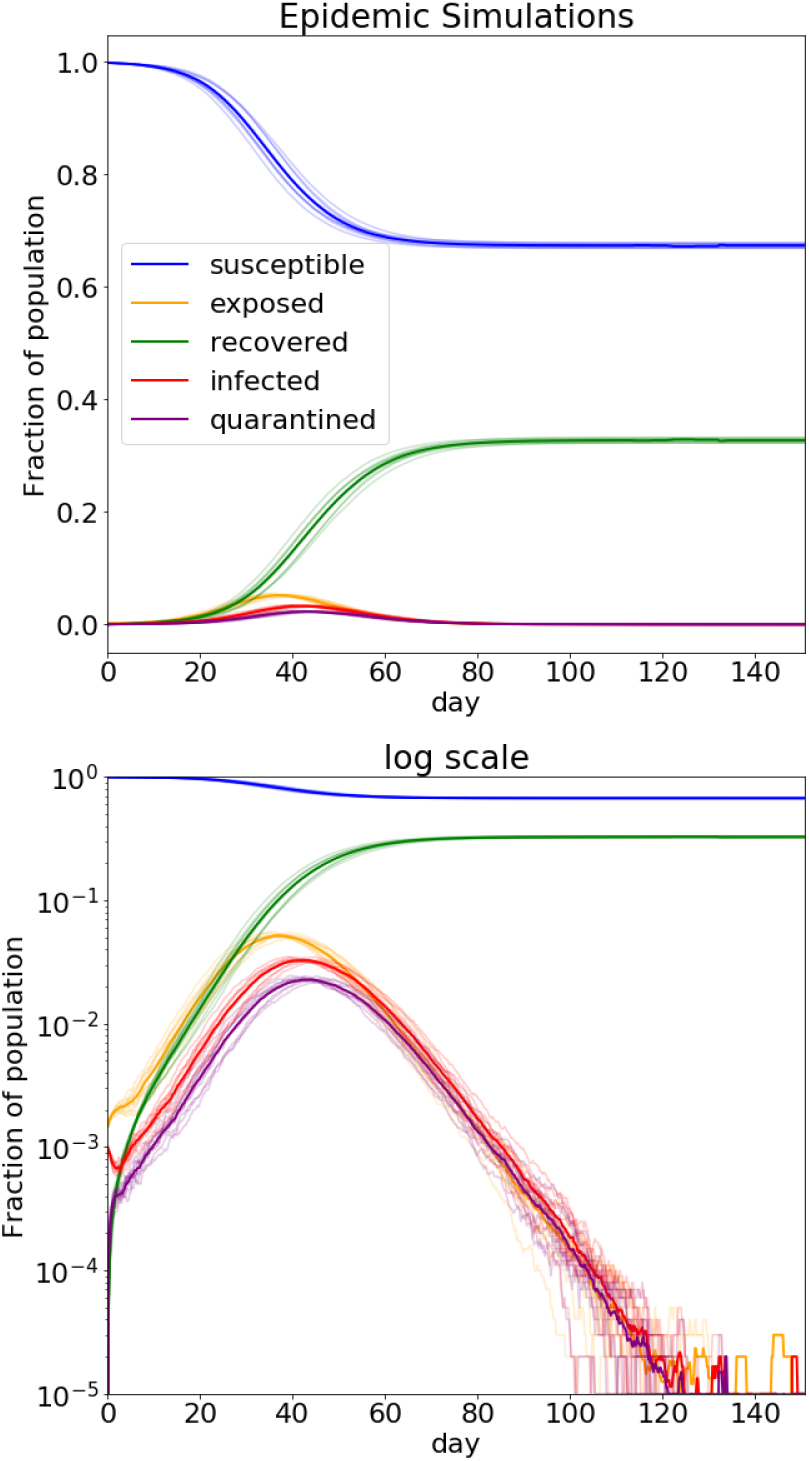
10 simulations with group sizes over time and their average (thick line). There is some spread of results, but mostly horizontal shifts in timing - aggregate outcomes are rather robust.

The strengths of this model are:

- It models quarantines and contact tracing.
- It models super-spreaders.
- By changing the graph structure, we will be able to model clustering and heterogeneity in Ro.
- This model can be flexibly adapted to changing parameters or policies.
- This model is open source.

The limitations of this model are:

- Many things are not modeled probabilistically, which should be: all neighbors are traced and quarantined, quarantined individuals never infect or get infected, tests come out positive for all Exposed and Infected.
- We do not model dynamic edges - nodes in contact stay in contact throughout.
- This is not the true graph form for human interactions, it is simplified. The real connection graph of people is more complex, with strong locality, but we do think it has a fat tailed degree distribution. We are unsure of the precise realistic parameters of the power law distribution, and results are sensitive to that. Parameters of the real-life graph (with and without social distancing) would be a valuable addition.
- The mean degree of nodes we selected is arbitrary (but we calibrate the infection probability to give the correct doubling time, so might correct that).
- We do not model asymptomatic patients directly. However, we capture them through the daily probability of recovery which was calculated to include them, and the probability an Infected node is detected which is not 1. So they are modeled as Infected who recover quickly and aren’t tested. This is an important avenue for future work since they might be very numerous.[1]
- We do not model superspreading events (gatherings, etc.) and these could be important if they increase the average number of interactions (and so infections) or if they mix the graph a lot.
- We do not model false negative test results, but these are equivalent to a smaller fraction of the population / Infected / neighbors being tested, so it is included in the probability of testing, which in fact represents the probability of detection. We also do not model false positive test results.

#### Parameter table

This is the parameter table for the simulation runs, along with their default values. Some of these were changed in the different simulations presented in the paper, as explained. Refer to the code for a full specification of all parameters and their use.

| parameter | value | meaning |
| --- | --- | --- |
| N | $10^5$ | Number of individuals in the simulation. |
| min_degree | 2 | Minimum degree of connection graph. |
| mean_degree | 20 | Mean degree of connection graph. |
| gamma | 0.2 | Parameter of degree distribution. Higher gamma means a fatter tail.<br>Default values make the top 0.1% degree = $15 \times$ median degree. |
| doubling_days | 3.1 | USA deaths data (worldometers.info): 11 -> 2,220 from March 4th to March 28th. |
| prob_infect | 0.022 | In a single interaction, what is the chance of an Infectious to infect a Susceptible.<br>Fitted to produce doubling_time For G with parameters $(10^5, 2, 20, 0.2)$. |
| prob_infect_exposed_factor | 0.5 | In a single interaction on a single day, how much more likely is an Exposed, in his infectious period before becoming contagious, to infect than an Infected.<br>$\text{prob\_infect\_exposed} = \text{prob\_infect} \times \text{prob\_infect\_exposed\_factor}$. |
| duration_exposed_infects | 2 | How many days before developing symptoms (becoming Infected) an Exposed individual is contagious. Considerable uncertainty. |
| incubation_duration_mean | 5.1 | Incubation period duration distribution mean. In days. Source. |
| incubation_duration_std | 4.38 | Incubation period duration distribution standard deviation. In days. Source. |
| prob_recover | 0.28 | Probability to change from Infections to Recovered in a single day. Inverse of expected recovery time. Source. |
| days_in_quarantine | 14 | Length of imposed quarantine, in days. |
| steps_per_day | 5 | Number of simulation steps per one day of real life time. |
| initial_infected_num | 100 | Number of individuals infected at $t=0$. |
| prob_infected_detected | * | Probability that an Infected individual is tested in a single day. |
| prob_neighbor_detected | * | Probability neighbor of a positive is traced and tested. |
| prob_exposed_detected | * | Probability Exposed individual tested in a single day.<br>This is the fraction of the general population tested daily. |

## Footnotes

1 See [popular article](https://www.cebm.net/covid-19/when-will-it-be-over-an-introduction-to-viral-reproduction-numbers-r0-and-re/)

2 See also this excellent implementation of a [similar model on GitHub](https://github.com/ryansmcgee/seirsplus).

3 This closed form formula of *P*_1_(*d*) is true in expectation, not for every specific second generation. It also assumes relatively few infectious nodes, so connections to multiple infectious nodes are neglected.

4 In fact, after the first step, this will be proportional to (*degree* − 1) since the node in question was infected by some other node, who can no longer be infected. We neglect this. To be even more precise, one would have to consider the clustering coefficient / locality of the graph to know how many neighboring nodes were not yet infected. Our graph is not local, so we do not address this.

5 As an aside we note that for a directed graph, a very similar argument gives 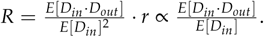

6 see [this preprint](https://cmmid.github.io/topics/covid19/tracing-bbc.html)

7 We estimated the doubling time from total number of reported COVID-19 deaths in the USA: from March 4th to March 28th, 2020 the number increased from 11 to 2220 deaths. the [Source](https://www.worldometers.info/coronavirus/country/us/). Similar numbers were observed in other geographies where no significant interventions were implemented.

## References

[1] Yan Bai, Lingsheng Yao, Tao Wei, Fei Tian, Dong-Yan Jin, Lijuan Chen, and Meiyun Wang. Presumed asymptomatic carrier transmission of COVID-19. JAMA, 323(14):1406, April 2020. doi: 10.1001/jama.2020.2565. URL https://doi.org/10.1001/jama.2020.2565.

[2] Martin Dottori and Gabriel Fabricius. Sir model on a dynamical network and the endemic state of an infectious disease. Physica A: Statistical Mechanics and its Applications, 434:25–35, 2015.

[3] John J Grefenstette, Shawn T Brown, Roni Rosenfeld, Jay De-Passe, Nathan TB Stone, Phillip C Cooley, William D Wheaton, Alona Fyshe, David D Galloway, Anuroop Sriram, Hasan Guclu, Thomas Abraham, and Donald S Burke. FRED (a framework for reconstructing epidemic dynamics): an open-source software system for modeling infectious diseases and control strategies using census-based populations. BMC Public Health, 13(1), October 2013.

[4] Joel Hellewell, Sam Abbott, Amy Gimma, Nikos I Bosse, Christopher I Jarvis, Timothy W Russell, James D Munday, Adam J Kucharski, W John Edmunds, Sebastian Funk, Rosalind M Eggo, Fiona Sun, Stefan Flasche, Billy J Quilty, Nicholas Davies, Yang Liu, Samuel Clifford, Petra Klepac, Mark Jit, Charlie Diamond, Hamish Gibbs, and Kevin van Zandvoort. Feasibility of controlling COVID-19 outbreaks by isolation of cases and contacts. The Lancet Global Health, 8(4):e488–e496, April 2020. doi: 10.1016/s2214-109x(20)30074-7. URL https://doi.org/10.1016/s2214-109x(20)30074-7.

[5] William Ogilvy Kermack and Anderson G McKendrick. A contribution to the mathematical theory of epidemics. Proceedings of the royal society of london. Series A, Containing papers of a mathematical and physical character, 115(772):700–721, 1927.

[6] Gary Wong, Wenjun Liu, Yingxia Liu, Boping Zhou, Yuhai Bi, and George F. Gao. MERS, SARS, and ebola: The role of super-spreaders in infectious disease. Cell Host & Microbe, 18(4):398–401, October 2015. doi: 10.1016/j.chom.2015.09.013. URL https://doi.org/10.1016/j.chom.2015.09.013.

